# Using HoloLens2™ to reduce staff exposure to aerosol generating procedures during a global pandemic

**DOI:** 10.1101/2020.05.24.20107193

**Authors:** John J Cafferkey, Dominic O P Hampson, Clare Ross, Angad S Kooner, Guy FJ Martin, James M Kinross

**Author notes:** Corresponding Author: Guy FJ Martin, telephone [withheld].

## Abstract

**RATIONALE:** COVID-19 poses a unique challenge; caring for patients with a novel, infectious disease whilst protecting staff. Some interventions used to give oxygen therapy are aerosol generating procedures. Staff delivering such interventions require PPE and are exposed to a significant viral load resulting in sick days and even death. We aim to reduce this risk using an augmented-reality communication device: The HoloLens by Microsoft.

**OBJECTIVES:** In a tertiary centre in London we aim to implement HoloLens technology, allowing other medical staff to remotely join the consulting clinician when in a high-risk patient area delivering oxygen therapy. The study primary outcome was to reduce the exposure to staff and demonstrate non-inferiority staff satisfaction when compared to not using the device. Our secondary outcome was to reduce extrapolated PPE costs when using the device.

**METHODS:** Our study was conducted in March and April 2020, within a respiratory unit delivering aerosolising oxygen therapies (High flow nasal oxygen, Continuous positive airway pressure and non-invasive ventilation) to patients with suspected or confirmed COVID-19 infection.

**MEASUREMENTS:** Self-reported questionnaires to assess satisfaction in key areas of patient care. An infrared people counting device was also used to assess staff in and out of the unit.

**MAIN RESULTS:** Mean self-reported time in the high-risk zone was less when using HoloLens (2.69 hours) compared to usual practice (3.96 hours) although this difference was not statistically significant (p = 0.3657). HoloLens showed non-inferiority when compared to usual practice in staff satisfaction score for all domains. Furthermore, mean staff satisfaction score encouragingly improved when using HoloLens. Self-reported PPE counts from this study showed 12 staff members changing PPE 25.8 times per shift, almost double the 13.5 times in the HoloLens count.

**CONCLUSIONS:** We have demonstrated HoloLens can reduce the number of staff exposed to aerosol generating areas in a novel infectious disease. Our results show it did not impair communication, medical staff availability or end of life care. HoloLens technology may also reduce the use of PPE, which has equipment availability and cost benefits. This study provides grounding for further use of the HoloLens device by bringing a bedside experience to experts remote to the situation.

## Text

COVID-19 is a highly transmissible viral pathogen spread through respiratory droplets or contact with contaminated surfaces(1). Contact with respiratory secretions is a known risk factor for infection in healthcare workers(2, 3). Non-invasive ventilation (NIV), continuous positive airway pressure (CPAP) and high flow nasal oxygen (HFNO) have been widely deployed in the treatment of COVID-19(4). These aerosol generating procedures (AGP) carry significant risk of airborne transmission via aerosolised virus to staff working in clinical areas where delivered.

Mixed reality (MR) technology merges real and virtual worlds, producing new environments and visualisations. HoloLens2™ is a MR head-mounted device developed and marketed by Microsoft Corporation (Redmond, WA, USA). Remote Assist 365 software permits bidirectional audio and visual communication for multiple users using the HoloLens2™ (Figure 1). Remote users see a first person view from the HoloLens2™ wearer, while the wearer experiences a holographic ‘heads up’ display, allowing them to see the their environment and relevant clinical data and imaging. The user interacts with these holographic objects using hand gestures or voice command, without the requirement for touch. HoloLens2™ has been deployed in medical education, surgery, and intensive care ward rounds (5, 6), but never before to protect staff from exposure to infectious disease in AGP areas.

**Figure 1.**
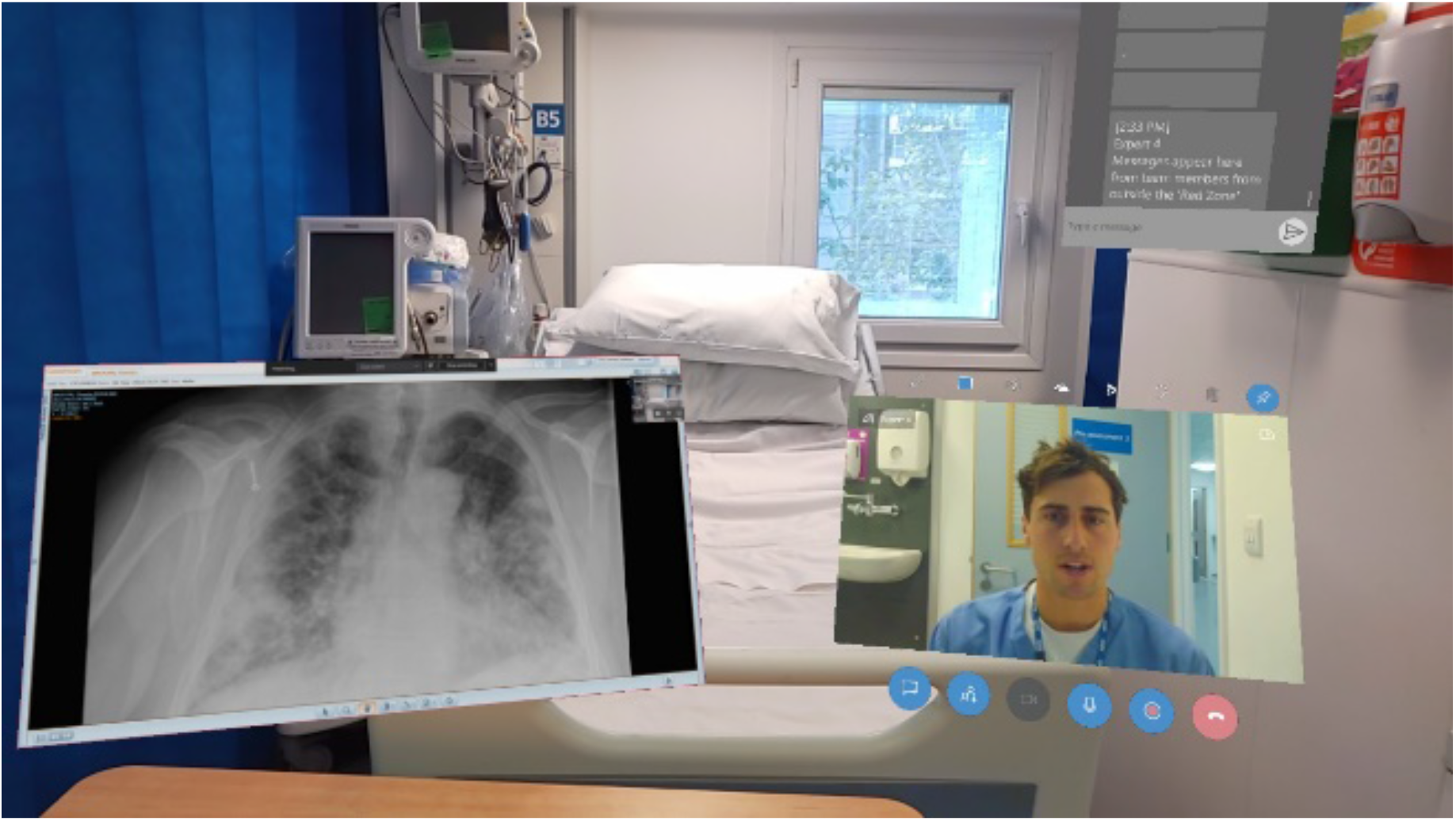
The “Red zone”. View of the HoloLens2™ user conducting the ward round. This is the view which is streamed to the junior team in the “clean area” during the ward round.

In the CPAP unit of a UK teaching hospital we implemented HoloLens2™ as a pragmatic technology-led improvement project to facilitate remote care during the COVID-19 pandemic in a “Red zone” where NIV/CPAP/HFNO is delivered. Pre-deployment practice was two or more staff donning personal protective equipment (PPE) for ward rounds and to review deteriorating patients. Following deployment of HoloLens2™, one doctor would conduct the ward round wearing the device and other team members would remain protected in the “clean” area whilst participating virtually in care delivery. Reviews of deteriorating patients by junior staff in the “Red zone” would also be conducted with HoloLens2™ to facilitate remote senior clinical input.

Evaluation of the intervention saw ward staff (doctors, nurses and allied health professionals) completing self-report questionnaires comparing pre- and post-HoloLens2™ deployment. Questions included time in “Red zone”, times donning /doffing and ten-point Likert scales assessing multiple domains of communication and care delivery. An infrared device counted people in and out of the “Red zone” (All-Tag, United Kingdom(7)/Sensor Development International, Netherlands(8)). Primary outcomes were: 1. Number of staff exposed to AGP environments before and after deployment of HoloLens2™, and 2. Self-reported safety and acceptability of HoloLens2™. A secondary outcome was reduction in PPE use. Local approval from Imperial College Healthcare NHS Trust as a technology led quality improvement project was obtained.

## Results

Over 15 days a total of 57 patients were cared for in the unit. The HoloLens2™ was used on 13 of the 15 days; a greater proportion than anticipated in large part reflecting the reluctance of the clinical team to revert to usual practice following deployment. The number of staff entering and leaving the “Red zone” was assessed for 36 hours of usual practice and 240 hours of HoloLens2™ use. Average “crossings” per hour of the antechamber were 12.5 and 10.3 respectively, a reduction of 17.6%.

21 questionnaires were completed. Average self-reported time in the “Red zone” was less when using HoloLens2™ (2.69 hours vs. 3.96 hours, p=0.3657). All domains assessed with Likert responses showed non-inferiority for care delivery when using the device. Furthermore, the mean satisfaction score for all domains increased during days using HoloLens2™. We found decreased self-reported antechamber use and an increase in confidence in the communicating critical clinical information (Table 1).

**Table 1.** Summary questionnaire responses from staff working in the CPAP ward on shifts where usual practice was observed or HoloLens2™ was used. All statistical tests are Mann Whitney U tests unless otherwise specified. *Unpaired t-test used.

| Domain | Usual Practice |  | HoloLens2™ |  | P-values |
| --- | --- | --- | --- | --- | --- |
|  | Mean | Median | Mean | Median |  |
| Time spent in PPE (hrs) | 3.96 | 3.00 | 2.69 | 1.25 | 0.3657* |
| Count time don/doff | 2.15 | 2.00 | 1.13 | 1.00 | 0.1139 |
| Antechamber use | n/a | n/a | n/a | n/a | 0.0082 |
| Communication | 7.62 | 8.00 | 8.50 | 8.00 | 0.8187 |
| Accessing medical support | 7.75 | 8.00 | 9.00 | 9.50 | 0.1494 |
| Communicating about sick patients | 7.08 | 7.00 | 9.14 | 9.00 | 0.0239 |
| Relaying information | 7.54 | 8.00 | 8.14 | 8.00 | 0.2874 |
| End of life care | 7.11 | 7.00 | 9.00 | 9.00 | 0.1793 |
| **n** | **8** |  | **13** |  |  |

For each healthcare professional joining the ward round remotely when using HoloLens2™ a full set of PPE is saved. Using self-reported donning/doffing counts, the use of HoloLens2™ led to a 48.7% reduction in the number of staff members donning/doffing each shift compared to usual practice (13.5 vs. 25.8). With this effect size, device purchase cost is covered in 212 days by PPE savings alone (9), and crucial PPE is saved at a time of international shortage.

## Discussion

This novel use of MR technology can be implemented rapidly to materially reduce staff exposure to high-risk environments. Critically, this does not impact quality of communication or staff perception of physician support. Our pilot data signals MR technology may improve confidence in clinical practice in high-risk areas. This objective improvement in communication is consistent with our subjective experience: first person, real-time audio and visual information allows senior medics to “get a feel” for patients they are consulting on despite physical distance.

An initial concern was device contamination, however, modifications to PPE protocols with infection prevention and control advice provided a satisfactory solution. We acknowledge that using HoloLens2™ in a novel disease with high PPE requirements may incur hidden risk, but on balance it is likely lower than exposing multiple additional staff to high-risk areas. Our results indicate a reduction in the use of PPE; clearly warranted during the global pandemic to protect resources. Furthermore, any PPE use is time consuming and risky; our intervention reduces its use.

A key weakness is the time-limited analysis of staff entering and leaving the “Red zone”, impeding our ability to objectively assess staff exposure to high-risk environments. However, our experience is that this reflects strong adoption of the HoloLens2^™^ by the clinical team. Whilst patients were universally accepting of the device, we acknowledge it does complicate critical non-verbal areas of communication like eye contact, and patient experience must be a measure of future implementation. Finally, lack of randomization and lack of control for patient numbers introduces further potential bias. During implementation, we needed to address concerns about data security, patient safety and cost. Our approach was necessarily pragmatic, rapidly delivering an intervention to protect staff during challenging times.

Our experience using the HoloLens2^™^ in high-risk clinical areas is promising, and reflects the ingenuity and flexibility needed during a pandemic. Widespread concern about a second wave of COVID-19 infection is compounded by the lack of an effective treatment and the inherent risk to healthcare workers. Further assessment will include objective measures of patient safety, possible contamination of devices and effectiveness in protecting healthcare workers.

## Data Availability

The data that support the findings of this study are available from the corresponding author, GM, upon reasonable request.

## Acknowledgments

We would like to thank Microsoft for their technological support during this project.Figure legends

## Footnotes

(no footnotes)

## Contributions

JC, DH, and AK devised and conducted the study. CR, GM and CR provided analysis advice, supervised the project and gave expert technical and medical support. All authors were involved in the write up and review of the manuscript.

## Support

Microsoft provided in-kind support with supply of devices and technical support. All Tag donated a two-way infrared people counter described below. Neither Microsoft nor All-Tag participated in the design, write up or review of the manuscript. The project was supported by the National Institute for Health Research (NIHR) Biomedical Research Centre based at Imperial College Healthcare NHS Trust and Imperial College London.

## Brief description

Mixed reality provides unique solutions to the unique challenges that COVID-19 presents to the health of staff caring for infected patients. We present a pilot study demonstrating the deployment of HoloLens2™ to reduce the number of medical staff exposed to high-risk clinical areas without compromising quality of care.

## Subject code

2.6 Information Technology/Informatics/Telemedicine

## MeSH terms

Telemedicine; COVID-19; Augmented Reality; Personal Protective Equipment;

## Notes

### Competing Interest Statement

The authors have declared no competing interest.

### Funding Statement

Microsoft provided in-kind support with supply of devices and technical support. All-Tag donated one two-way infrared people counter described below. Neither Microsoft nor All-Tag participated in the design, write up or review of the manuscript. The project was supported by the National Institute for Health Research (NIHR) Biomedical Research Centre based at Imperial College Healthcare NHS Trust and Imperial College London.

### Author Declarations

The project had local institutional approval as a technology-led quality improvement project from Imperial College Healthcare NHS Trust.

